# A COMPARATIVE EVALUATION OF NON-INCISED PAPILLAE AND ENTIRE PAPILLA PRESERVATION SURGICAL APPROACHES IN TREATMENT OF INTRABONY DEFECTS – A CLINICAL AND RADIOGRAPHIC STUDY

**DOI:** 10.1101/2022.03.27.22273001

**Authors:** Bandi Divya Elenora Martina, Rampalli Viswa Chandra, Vallabhdas Santosh, Gollapalle Prabhandh Reddy, Aileni Amarender Reddy

## Abstract

**Aim & Objectives:** The aim of the present study was to clinically and radiographically compare and evaluate the NIPSA (Non-Incised Papillae Surgical Approach) and EPP (Entire Papilla Preservation) techniques in the treatment of intra bony defects.

**Patients and methods:** From this initial patient pool of 156 patients, 44 individuals satisfying the inclusion criteria were selected. One site in each subject was assigned into each of the following experimental groups which were treated with the relevant procedure; NIPSA and EPP groups. Clinical parameters included the recording of pocket probing depths (PPD), clinical attachment levels (CAL) and papilla loss. The evaluation of bone fill was performed at the end of 3 & 6-months by using Image J® software.

**Results:** NIPSA and EPP resulted in highly significant CAL gains (4.00±1.00 *vs* 5.25 ± 1.47mm; *p*≤*0*.*001)* and PPD reductions (3.352±0.70 *vs* 3.625±1.024mm; *p*≤*0*.*001)* at 6-months post-surgery. From the start to the end of study period, NIPSA resulted in almost no papilla loss (1.71±0.47 to 1.73±0.77mm) while EPP technique showed minimal and insignificant (1.57±0.52 to 1.48±0.71mm; *p*≥*0*.*05)* papilla loss. The EPP group revealed a highly significant (0.459±0.16 *vs* 0.269±0.16cm^*2;*^ *p*≤*0*.*001)* bone fill over NIPSA group at 6-months.

**Conclusion:** From this trial conducted over a period of 6-months, NIPSA and EPP both resulted in significant improvements in clinical outcomes. NIPSA and EPP showed favourable outcomes in terms papilla loss and bone fill respectively. Both the techniques achieve the aims outlined earlier and broaden the choice available to a periodontist in the management of intrabony defects.

## INTRODUCTION

Intraosseous defects are a frequent sequela of periodontitis which when left untreated may significantly affect the long-term success of the defect associated teeth.^1,2^ The morphology of the defect needs to be carefully considered when selecting the regenerative approach as the potential for regeneration depends on the number of osseous walls surrounding the defect.^3-6^ However, tissue shrinkage and loss with exposure and contamination of the graft or regenerative material has been associated with reduced clinical outcomes.^7,8^

Conventional periodontal surgery results in pocket probing depth reduction and elimination of osseous defects^6-8^ and may also be associated with considerable loss of the tooth’s supporting tissue including the papilla.^9,10-13^ To offset problems related to conventional periodontal surgery such as papillae loss, postoperative pain, bleeding, swelling, root hypersensitivity and delayed healing,^11-13^ several papilla preservation techniques^14-16^ have been employed to increase the rate of clinical success in periodontal surgery. The adoption of these approaches improves the degree of primary wound closure and results in optimum stabilization of the wound.^17-20^

The EPP (Entire Papilla Preservation) technique has recently been employed for regenerative treatment of isolated deep intrabony defects.^21^ While providing adequate access for debridement, the basic principle of this technique is to preserve the full integrity of the defect-associated papilla that provides an intact gingival chamber over the intrabony defect, and maintains vascular integrity of the defect associated papilla.^21,22^ Similarly, an apical incision approach termed as NIPSA (Non-Incised Papillae Surgical Approach) was employed, without incisions or tear of interdental papillae or marginal tissues for the treatment of the periodontal defects.^23,24^ The techniques promise minimal flap manipulation, optimum stabilization of the flap, inter-dental tissue preservation and adequate support to regenerative materials.^21-25^

In light of the reported advantages of these procedures, this study aimed to compare the effectiveness of the NIPSA (Non-Incised Papillae Surgical Approach) and EPP (Entire Papilla Preservation) techniques in the treatment of intra bony defects. The evaluation was made post-operatively based on improvements of select outcomes such as pocket probing depths (PPD), clinical attachment levels (CAL), papilla loss and bone fill.

## MATERIALS AND METHODS

### Sample Size Calculation and Study methodology

To have an 80% chance (β error) of detecting a significant (two-sided 5% level) and a largest difference of 1mm in probing depth between groups with a standard deviation of 0.1, 17 subjects per group were required.

A non-randomised protocol was followed where two calibrated investigators (DEMB &VS) selected subjects (30-60 years) with chronic periodontal disease from the outpatient section of the Department of Periodontology with two closely matched sites in different arches with i) presence of two or more intrabony defects (ii) average probing pocket depth (PPD) between 5-8mm, (iii) average clinical attachment level (CAL) between 5-10mm, and with (iv) radiographic depth≥4mm. Exclusion criteria comprised of (1) Defects involving lingual sites, (2) patients with a history of systemic conditions that affect the periodontium, and (3) smokers. From this initial patient pool of 156 patients, 44 individuals satisfying the inclusion criteria were selected with all the participants providing informed consent and the study protocol was approved by the institutional ethics committee (SVSIDS/PERIO/2/2018).

### Outcomes

The following clinical parameters^26,27^ were recorded at the time of the surgery, 3 & 6 months; *(1)* Clinical attachment level (CAL), measured from the cementoenamel junction (CEJ) to the base of the pocket (*2)* Probing pocket depth (PPD) measured from the gingival margin to the base of the pocket *(3)* Papilla height^27^ was calculated by measuring the distance between the papilla tip (PT) and the contact point (CP) between the two adjacent teeth. Radiographically, bone regeneration^26^ was assessed from baseline to 3 months and 6months as follows; The radiograph obtained at 3 and 6 months were subtracted from the radiograph taken at the baseline by using commercially available image processing software (Adobe Photoshop® 6.0, Adobe Systems, San Jose, USA). To reduce the brightness and contrast variations, both images were adjusted based on the levels and the curves in the software. Before digital subtraction, both radiographs were moved in appropriate directions as needed, to reduce geometric distortion. These images were then superimposed and subtracted by selecting the image≥calculation≥exclusion≥new channel tools. The excluded residual bone height was outlined by using the polygonal lasso tool and the layer was copied and saved as a separate joint photographic expert group (JPEG) document at low compression.^26^

After digital subtraction, the digitized and excluded residual bone height was transferred to open-source software for area calculation (Image J®, Research Services Branch, NIH, Bethesda, Maryland, USA) for area calculation. The layer was converted into a grayscale image, and the measurement scale was set to account for any magnification/reduction of the radiograph because of the RVG. The area of the layer was calculated *(Cm*^*2*^) by initially enclosing the entire area with the rectangular selection tool and then by using Analyze≥Analyze Particles tool.^26^

### Operative Procedures

#### Study protocol

After the interdental areas were probed buccally and lingually/palatally, the site was considered for the study if the deepest probing depth (PPD) at the buccal site was ≥5mm, not involving predominant lingual defects. All baseline (on the day of surgery) parameters were recorded before the surgical procedure and at 3 and 6-months. Clinical parameters were recorded using a University of North Carolina no.15 (UNC-15) color-coded periodontal probe.

#### NIPSA group

After mapping of the limit of the defect, a buccal horizontal or oblique incision was placed on cortical bone, as apically as possible while preserving the suprabony soft tissue to protect the defect. Mesodistally, the incision was extended to across the entire length of the defect. The soft tissue was reflected apicocoronally to expose the bone crest, maintaining the marginal tissue integrity and papillae architecture. The granulation tissue and epithelium of the pocket were eliminated with mini-curettes while respecting the marginal soft tissue and residual fibers attached to the cementum. The affected root was scaled and planed, and calculus eliminated with ultrasonic tips. Once the defect was debrided, the regenerative materials were applied and the incision line was sutured.

### EPP group

After administration of local anaesthesia, a buccal intra-crevicular incision of the defect-associated tooth was performed. A short vertical incision was later extended in the buccal gingiva contralateral to the osseous defect and extended beyond the mucogingival line. Following elevation of a buccal full-thickness mucoperiosteal flap extending from the vertical incision to the defect-associated papilla, periosteal elevator facilitated an undermining tunnel preparation of the defect associated papilla. Outmost care was taken to elevate the interdental papilla in a full-thickness manner to the intact lingual bone crest’s coronal edge. Granulation tissue was removed from the inner aspect of the defect-associated interdental papilla with surgical scissors. Any remaining granulation tissue was removed with a mini-curette. An ultrasonic scaler was used to remove any residual subgingival plaque or calculus from the exposed root surface. Once the defect was debrided, the regenerative materials were applied, and the incision line was sutured.

#### Application of biomaterial

The application of the biomaterial was identical in the two groups. After defect debridement and instrumentation of the root surface, tetracycline was applied to the root surface for 2 minutes to remove the smear layer, followed by using hydroxyapatite graft (G-graft®, Saharanpur, UP, India). Finally, a GTR membrane (Pro-Tiss®, AzureBio, Madrid, Spain) was placed.

### Statistical Analysis

Data was analyzed by using Prism8® (GraphPad Software, La Jolla, USA). Intragroup comparison was performed by using ANOVA followed by multiple comparisons using Bonferroni correction. One-way ANOVA followed by the post hoc test was used for intragroup and intergroup comparisons. A p≤0.001 was considered as highly significant, p≤0.05 as significant and p>0.05 as non-significant.

## RESULTS

All subjects (Age:43.99±12.27; 18 males) completed the study. 9 subjects were lost during follow up to various reasons and the final statistical analysis was limited to 35 subjects.

### Intragroup Comparisons

#### Probing Pocket Depth

The mean probing pocket depths (in *mm*) in the NIPSA and EPP groups were 8.235±1.52, 5.117±0.69, 3.352±0.70 and 8.00±1.15, 5.062±1.123, 3.625±1.024 at baseline and at the end of 3 & 6 months respectively *(Table 1)*. The intragroup reduction when compared from baseline to 3 & 6 months was statistically highly significant in both the treatment groups *(p*≤*0*.*001)*.

**Table 1:** Intergroup comparison of PPD & CAL (in *mm*) at different time-intervals.

| Variable | Interval | NIPSA | EPP | t-value | p-value |
| --- | --- | --- | --- | --- | --- |
| **PPD** | *Baseline* | 8.235±1.52 | 8.00±1.15 | 0.497 | 0.311(NS) |
|  | *3-months* | 5.117±0.69 | 5.062±1.123 | 0.170 | 0.432(NS) |
|  | *6-months* | 3.352±0.70 | 3.625±1.024 | -0.899 | 0.188(NS) |
| **CAL** | *Baseline* | 9.53±1.37 | 8.69±1.45 | 1.71 | 0.09(NS) |
|  | *3-months* | 6.12±0.70 | 6.69±1.40 | 1.40 | 0.14(NS) |
|  | *6-months* | 4.00±1.00 | 5.25±1.44 | 2.91 | 0.0066* |

*Independent t-test, $p \leq 0.05$ \*(significant), $p \leq 0.001$ \*\* (highly significant), $p \geq 0.05$ NS (Not significant).*

#### Clinical attachment level

The mean clinical attachment level (in *mm*) in NIPSA and EPP groups were 9.529±1.37, 6.117±0.69, 4.00±1.00 and 8.687±1.44, 6.687±1.30, 5.25±1.47 at baseline and at the end of 3 & 6 months respectively. This intragroup reduction from baseline to 3 & 6 months was statistically highly significant in both the treatment groups *(p*≤*0*.*001)*.

#### Papilla Loss

From the start to the end of study period, NIPSA resulted in almost no papilla loss (1.71±0.47 to 1.73±0.77mm) while EPP technique showed minimal and insignificant (1.57±0.52 to 1.48±0.71mm; *p*≥*0*.*05)* papilla loss.

#### Bone Fill

The bone fill *(cm*^*2*^) when compared from baseline to 3 & 6 months in the NIPSA and EPP groups were 0.244±0.12 and 0.269±0.16 and 0.484±0.16 and 0.459±0.16 respectively. This intragroup bone fill compared from baseline to end of 3 months and from baseline to 6 months was statistically significant in both the treatment groups (p≤0.05).

### Intergroup Comparisons

No significant differences were observed between the groups for PPDs at different time intervals. At 6 months, there was a substantial difference in CAL between the groups *(p*≤*0*.*05) (Table 1)*. Recordings of papilla loss revealed no significant difference from baseline to 3 & 6 months between the treatment groups *(Table 2)*. When compared from baseline to 3 & 6 months, the intergroup bone fill was statistically highly significant *(p*≤*0*.*001)* in between the groups. The EPP group revealed a highly significant *(p*≤*0*.*001)* and higher bone fill over the NIPSA group at 3 and 6 months *(Table 3)*.

**Table 2:** Intergroup comparison of papilla loss at various time-intervals.

| Time | Groups | Mean and standard deviation | Mean difference | Std error | t-value | p-value | 95%confidence interval |  |
| --- | --- | --- | --- | --- | --- | --- | --- | --- |
|  |  |  |  |  |  |  | Lower bound | Upper bound |
| **Baseline** | *NIPSA* | 1.71±0.47 | 0.21 | 0.172 | 1.199 | 0.24 (NS) | -0.14 | 0.56 |
|  | *EPP* | 1.57±0.52 |  |  |  |  |  |  |
| **3-months** | *NIPSA* | 1.69±0.78 | 0.27 | 0.17 | 1.57 | 0.13 (NS) | 0.08 | 0.62 |
|  | *EPP* | 1.44±0.67 |  |  |  |  |  |  |
| **6-months** | *NIPSA* | 1.73±0.77 | 0.27 | 0.17 | 1.57 | 0.13 (NS) | 0.08 | 0.62 |
|  | *EPP* | 1.48±0.71 |  |  |  |  |  |  |

*Independent t-test, $p \leq 0.05$ \*(significant), $p \leq 0.001$ \*\* (highly significant), $p \geq 0.05$ NS (Not significant).*

**Table 3:** Intergroup comparison of bone fill *(Cm2)* at baseline to 3 months and baseline to 6 months.

| Groups | NIPSA | EPP |
| --- | --- | --- |
| **Baseline to 3-months** | 0.244±0.12 | 0.484±0.16 |
| **Baseline to 6-months** | 0.269±0.16 | 0.459±0.16 |
| **t-value** | -4.65 | -3.409 |
| **p-value** | $\leq 0.001$** | $\leq 0.001$** |

*Independent t-test, $p \leq 0.05$ \*(significant), $p \leq 0.001$ \*\* (highly significant), $p \geq 0.05$ NS (Not significant).*

## DISCUSSION

The modern periodontal paradigm is to preserve the inter-dental papillae through an adequate design of the flap and to achieve primary wound closure following regenerative surgery in intrabony defects.^14,29^ The flap design represents an essential parameter in any surgical technique in which the marginal tissue stability and good re-vascularization during early wound healing^30,31^ is crucial in promoting a stable closure of the wound. The aim of the present study was to clinically and radiographically compare and evaluate the NIPSA and EPP techniques in the treatment of intra bony defects.

Liu et al^32^ conducted a meta-analysis on minimally invasive surgery with and without regenerative materials and stated that there was no significant difference in clinical outcomes between the groups. Previous reports investigating the use of different minimally invasive techniques in association with biomaterials,^19,31^ reported a mean CAL gain of 4.8 mm and a mean PD reduction of 5.2 mm. Similar results were seen in our study with CAL gains and PD reduction of 5.53mm & 4.88mm and 3.44mm & 4.38mm mm in NIPSA and EPP groups respectively when regenerative biomaterials were used. Moreno Rodriguez et al^24^ compared a minimally-invasive surgical technique (MIST) and a non-incised papillae surgical approach (NIPSA) and showed significant improvement in CAG in NIPSA than MIST.^24^ These results are comparable with the data reported in previous studies based on the concepts of minimally invasive surgery.^19,31,32^ Similarly, the present study revealed no significant differences in PPD but revealed a significant change in CAG *(p*≤*0*.*05)* with more favorable results in NIPSA group at six months. Differences in CAL gain between the groups may be attributed, at least in part, to differences in the status of disease (chronic/aggressive periodontitis) of the study population, protocols for postoperative maintenance, follow-up intervals, and flap design and soft tissue management. In the case of NIPSA, the incision is placed dissecting the subperiosteal gingival vessels near the mucogingival junction,^33^ thereby maintaining the marginal integrity. The non-incised gingival vessels show continuity with those of the periodontal ligament that minimizes surgical trauma in marginal tissues.

Ruiz and Caffese^24^ revealed a significant papillae recession in MIST compared with no recession in NIPSA. NIPSA has an added advantage as it is not reflected at the intrasulcular level, minimizing the post-surgical gingival recession. Velvart et al^34^ investigated the papillary shrinkage after sulcular incision in patients with healthy periodontium, and reported a gradual increase in the papilla loss during healing. In our study, papilla loss recordings remained at the same values during follow-up in NIPSA; but, a very limited, but not significant, amount of papilla loss was observed in EPP. When the papilla is not elevated, the supracrestal fibers connecting the interdental tissues to the crest associated tooth are not dissected and this keeps the papilla in its coronal position. This helps maintain space under the papilla resulting in the post-surgical stability of the interproximal soft tissues. In EPP group, intracrevicular incision given may result in the retraction of the papilla.

Previous studies on different papilla preservation techniques also revealed significant radiographic outcomes.^35-38^ On the contrary, the present investigation revealed significant differences in bone fill of intrabony defects in the radiographic outcomes following treatment with NIPSA or EPP with the application of HA/GTR. This change may be attributed to the selection of sites and the type of defect. In the case of NIPSA, it was performed in multiple intraosseous defects with defect configuration, including one and/or two wall components, always involving the buccal wall. In contrast, EPP was performed in isolated one or two intrabony defects with 2-wall components with a missing buccal wall and a relatively well-preserved lingual wall. Minimal wound dehiscences are seen^31^ and papilla preservation techniques result in uneventful primary closure.^19,39^ EPP showed mild papillary loss and this may be because of the proximity of the initial incision to the papilla. Von Arx et al^40^ stated that the type of incision technique is crucial as it affects the healing of marginal tissues. Previous studies^23,24^ have stated that defects treated with NIPSA showed lesser inflammation than other techniques post-operatively. This may have contributed to better clot stability and greater bone-fill in NIPSA over EPP.^19,31,39,40^

From this trial conducted over a period of 6-months, NIPSA and EPP both resulted in significant improvements in clinical outcomes. NIPSA and EPP showed favourable outcomes in terms papilla loss and bone fill respectively. Both the techniques achieve the aims outlined earlier and broaden the choice available to a periodontist for papilla preservation in the management of intrabony defects.

## Data Availability

All data produced in the present work are contained in the manuscript

## Figure Legends

**Figure 1:**
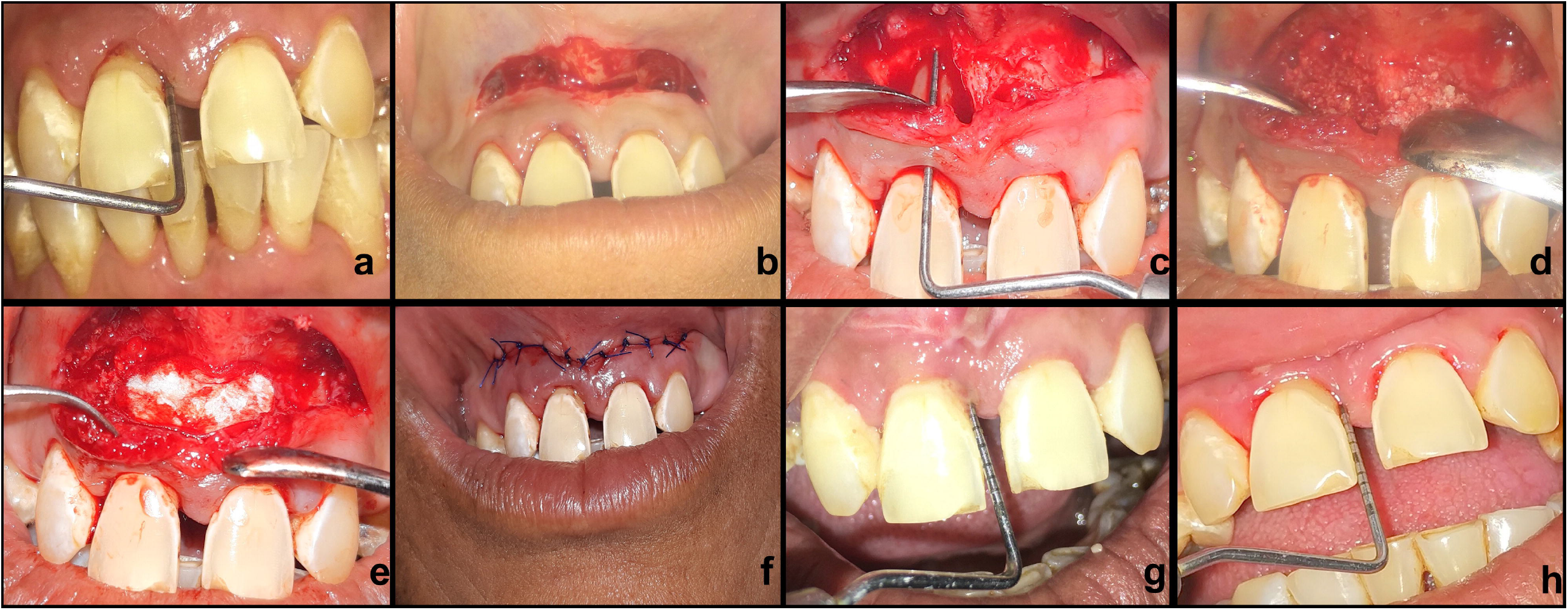
NIPSA technique; Pre surgical probing *(a)*. A buccal horizontal or oblique incision was placed correctly on cortical bone, as apically as possible and the incision was extended mesiodistally to access the defect *(b)*. The soft tissue is reflected apicocoronally to expose the bone crest, maintaining the marginal tissue integrity and papillae architecture *(c)*. Once the defect was debrided, the regenerative materials *(d, e)* were applied, and the incision line was sutured *(f)*. The sites were probed again at 3*(g)* and 6 months *(h)*.

**Figure 2:**
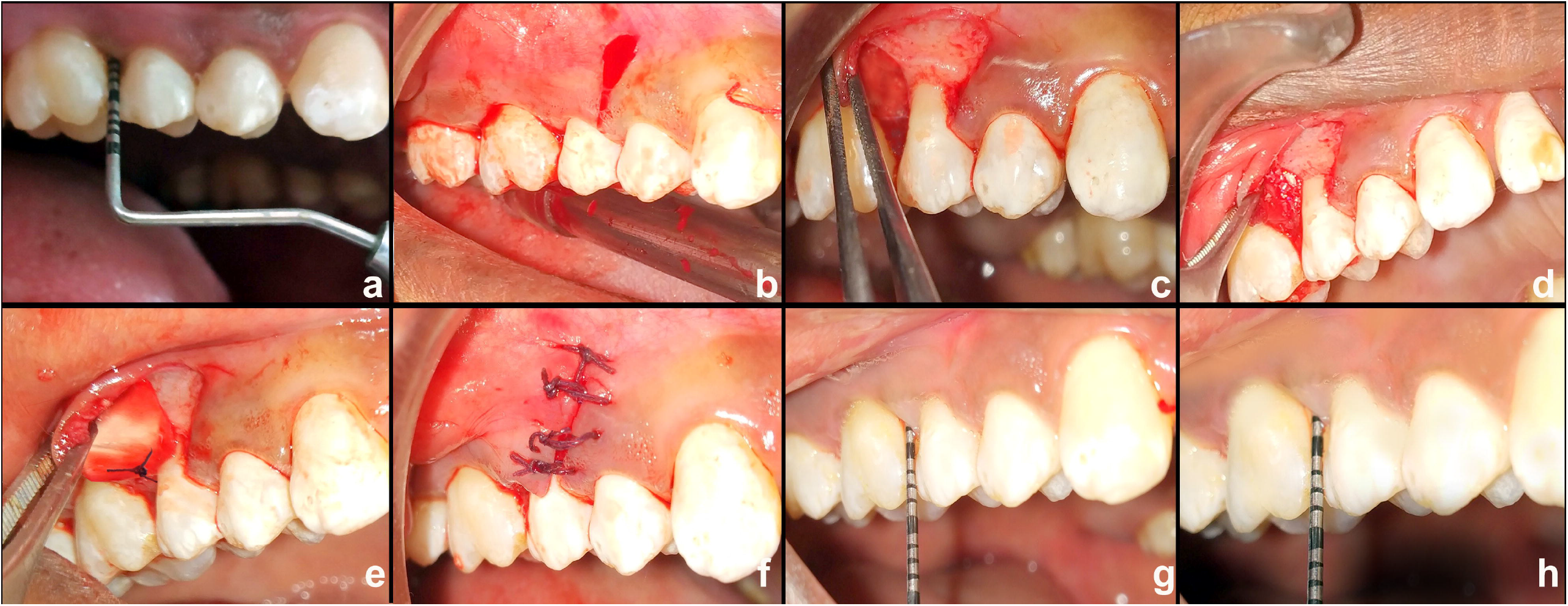
EPP Technique; Pre surgical probing *(a)*. A buccal intra-crevicular incision was given at the defect-associated tooth. A short vertical incision was later extended in the buccal gingiva contralateral to the osseous defect and extended beyond the mucogingival line *(b)*. Following the elevation of a buccal full-thickness mucoperiosteal flap extending from the vertical incision to the defect-associated papilla *(c)*, periosteal elevator instrument facilitated the undermining tunnel preparation of the defect associated papilla. Once the defect was debrided, the regenerative materials were applied *(d, e)*, and the incision line was sutured *(f)*. The sites were probed again at 3*(g)* and 6 months *(h)*.

**Figure 3:**
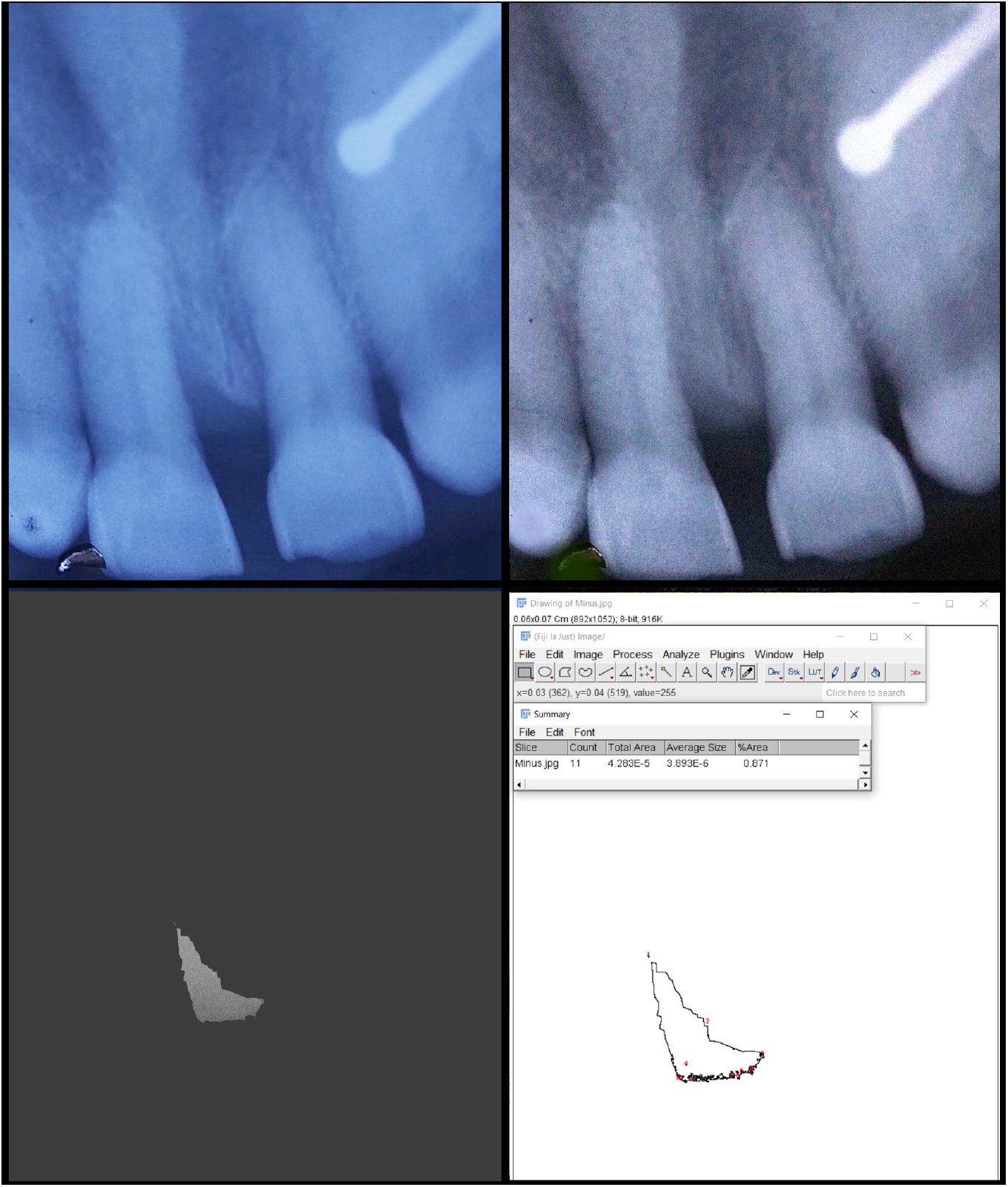
The initial radiographic image obtained at baseline *(top-left)* was subtracted from the radiographic images taken at 3 or 6-months *(top-right)*, in Adobe Photoshop® 6.0. The obtained layer *(bottom-left)* was transferred into Image J® software for area calculation *(bottom-right)*.

## REFERENCES

1. Papapanou PN, Tonetti MS. Diagnosis and epidemiology of periodontal osseous lesions. Periodontol 2000 2000;22(1):8–21.

2. Hägi TT, Laugisch O, Ivanovic A, Sculean A. Regenerative periodontal therapy. Quintessence Int 2014;45(3):185–92.

3. Cortellini P, Prato GP, Tonetti MS. Long□term stability of clinical attachment following guided tissue regeneration and conventional therapy. J Clinical Periodontol 1996;23(2):106–11.

4. Cortellini P, PiniCPrato G, Tonetti M. Periodontal regeneration of infrabony defects (V). Effect of oral hygiene on longCterm stability. J Clinical Periodontol 1994;21(9):606–10.

5. Cortellini P, Prato GP, Tonetti MS. The modified papilla preservation technique. A new surgical approach for interproximal regenerative procedures. J Periodontol 1995;66(4):261–6.

6. Cortellini P, Prato GP, Tonetti MS. The simplified papilla preservation flap. A novel surgical approach for the management of soft tissues in regenerative procedures. Int J Periodontics Restorative Dent 1999;19(6).

7. Tonetti MS, Cortellini P, Suvan JE, Adriaens P, Baldi C, Dubravec D, Fonzar A, Fourmousis I, Magnani C, Muller□Campanile V, Patroni S. Generalizability of the added benefits of guided tissue regeneration in the treatment of deep intrabony defects. Evaluation in a multi□center randomized controlled clinical trial. J Periodontol 1998;69(11):1183–92.

8. Cortellini P, Tonetti MS, Lang NP, Suvan JE, Zucchelli G, Vangsted T, Silvestri M, Rossi R, McClain P, Fonzar A, Dubravec D. The simplified papilla preservation flap in the regenerative treatment of deep intrabony defects: clinical outcomes and postoperative morbidity. J Periodontol 2001;72(12):1702–12.

9. Polson AM, Heul LC. Osseous repair in infrabony periodontal defects. J Clinical Periodontol 1978;5(1):13–23.

10. Kaldahl WB, Kalkwarf KL, Patil KD, Molvar MP, Dyer JK. Long□term evaluation of periodontal therapy: I. Response to 4 therapeutic modalities. J Periodontol 1996;67(2):93–102.

11. Bosshardt DD, Sculean A. Does periodontal tissue regeneration really work? Periodontol 2000 2009;51(1):208–19.

12. Sculean A, Alessandri R, Miron R, Salvi GE, Bosshardt DD. Enamel matrix proteins and periodontal wound healing and regeneration. Clin Adv Periodontics 2011;1(2):101–17.

13. Stoecklin-Wasmer C, Rutjes AW, Da Costa BR, Salvi GE, Jüni P, Sculean A. Absorbable collagen membranes for periodontal regeneration: a systematic review. J Dent Res 2013;92(9):773–81.

14. Takei HH, Han TJ, Carranza Jr FA, Kenney EB, Lekovic V. Flap technique for periodontal bone implants: Papilla preservation technique. J Periodontol 1985;56(4):204–10.

15. Cortellini P, Prato GP, Tonetti MS. The modified papilla preservation technique. A new surgical approach for interproximal regenerative procedures. J Periodontol 1995;66(4):261–6.

16. Cortellini P, Prato GP, Tonetti MS. The simplified papilla preservation flap. A novel surgical approach for the management of soft tissues in regenerative procedures. Int J Periodontics Restorative Dent 1999;19(6).

17. Tibbetts LS, Shanelec D. Periodontal microsurgery. Dent Clin North Am 1998;42(2):339.

18. Cortellini P, Tonetti MS. Microsurgical approach to periodontal regeneration. Initial evaluation in a case cohort. J Periodontol 2001;72(4):559–69.

19. Cortellini P, Tonetti MS. A minimally invasive surgical technique with an enamel matrix derivative in the regenerative treatment of intra□bony defects: A novel approach to limit morbidity. J Clinical Periodontol 2007;34(1):87–93.

20. Cortellini P, Tonetti MS. Improved wound stability with a modified minimally invasive surgical technique in the regenerative treatment of isolated interdental intrabony defects. J Clinical Periodontol 2009;36(2):157–63.

21. Aslan S, Buduneli N, & Cortellini P. (2017a). Entire papilla preservation technique: A novel surgical approach for regenerative treatment of deep and wide intrabony defects. Int J Periodontics Restorative Dent 2017:37, 227–233.

22. Aslan S, Buduneli N, Cortellini P. Entire papilla preservation technique in the regenerative treatment of deep intrabony defects: 1□Year results. J Clinical Periodontol 2017;44(9):926–32.

23. Rodriguez JA, Caffesse RG. Nonincised Papillae Surgical Approach (NIPSA) in Periodontal Regeneration: Preliminary Results of a Case Series. Int J Periodontics Restorative Dent 2018;38(Suppl):s105-s111.

24. Moreno Rodríguez JA, Ortiz Ruiz AJ, Caffesse RG. Periodontal reconstructive surgery of deep intraosseous defects using an apical approach. Non□incised papillae surgical approach (NIPSA): A retrospective cohort study. J Periodontol 2019;90(5):454–64.

25. Sculean A, Gruber R, Bosshardt DD. Soft tissue wound healing around teeth and dental implants. J Clin Periodontol 2014 ;41:S6–22.

26. Soni R, Priya A, Yadav H, Mishra N, Kumar L. Bone augmentation with sticky bone and platelet-rich fibrin by ridge-split technique and nasal floor engagement for immediate loading of dental implant after extracting impacted canine. Nat J Maxillofacial Surg 2019;10(1):98.

27. Nordland WP, Tarnow DP. A classification system for loss of papillary height. J Periodontol 1998;69(10):1124–6.

28. Poulsen S. Epidemiology and indices of gingival and periodontal disease. Pediatr Dent 1981;3:82–8.

29. Evian CI, Corn H, Rosenberg ES. Retained interdental papilla procedure for maintaining anterior esthetics. Compend Contin Educ Dent 1985;6(1):58.

30. Tonetti MS, Lang NP, Cortellini P, Suvan JE, Adriaens P, Dubravec D, Fonzar A, Fourmousis I, Mayfield L, Rossi R, Silvestri M. Enamel matrix proteins in the regenerative therapy of deep intrabony defects: A multicentre randomized controlled clinical trial. J Clinical Periodontol 2002;29(4):317–25.

31. Mörmann W, Ciancio SG. Blood supply of human gingiva following periodontal surgery. A fluorescein angiographic study. J Periodontol 1977;48(11):681–92.

32. Liu S, Hu B, Zhang Y, Li W, Song J. Minimally invasive surgery combined with regenerative biomaterials in treating intra-bony defects: a meta-analysis. PloS one 2016;11(1):e0147001.

33. Nakayama Y, Soeda S, Kasai Y. The importance of arterial inflow in the distal side of a flap: An experimental investigation. Plast Reconstr Surg 1982;69(1):61–7.

34. Velvart P, Ebner-Zimmermann U, Ebner JP. Papilla healing following sulcular full thickness flap in endodontic surgery. Oral Surg Oral Med Oral Pathol Oral Radiol Endod 2004;98(3):365–9.

35. Steffensen B, Weber HP. Relationship between the radiographic periodontal defect angle and healing after treatment. J Periodontol 1989;60(5):248–54.

36. Kuriakose A, Majo Ambooken JJ, John P. Modified Whale’s tail technique for the management of bone-defect in anterior teeth. J Ind Soc Periodontol 2015 ;19(1):103.

37. Di Tullio M, Femminella B, Pilloni A, Romano L, D’Arcangelo C, De Ninis P, Paolantonio M. Treatment of supra□alveolar□type defects by a simplified papilla preservation technique for access flap surgery with or without enamel matrix proteins. J Periodontol 2013;84(8):1100–10.

38. Fickl S, Thalmair T, Kebschull M, Böhm S, Wachtel H. Microsurgical access flap in conjunction with enamel matrix derivative for the treatment of intra□bony defects: A controlled clinical trial. J Clinical Periodontol 2009;36(9):784–90.

39. Aslan S, Buduneli N, Cortellini P. Clinical outcomes of the entire papilla preservation technique with and without biomaterials in the treatment of isolated intrabony defects: A randomized controlled clinical trial. J Clinical Periodontol 2020 ;47(4):470–8.

40. Von Arx T, AlSaeed M, Salvi GE. Five-year changes in periodontal parameters after apical surgery. J Endod 2011 Jul 1;37(7):910–8.

